# Fifteen-year Alcohol Consumption Trajectories and Their Association with Cardiovascular Events and Mortality: the Framingham Heart Study

**DOI:** 10.1101/2024.11.09.24317038

**Authors:** Yuanming Leng, Huitong Ding, Yi Li, Xue Liu, Mengyao Wang, Chenglin Lyu, Daniel Levy, Jiantao Ma, Chunyu Liu

## Abstract

**Background:** Long-term alcohol consumption patterns play a critical role in human health.

**Objectives:** We aimed to identify alcohol consumption trajectories and analyze their associations with coronary heart disease (CHD) and all-cause mortality in the Framingham Heart Study (FHS).

**Methods:** We used a growth mixture model to identify sex-specific alcohol consumption trajectories over 15 years and Cox regression to examine associations of these trajectories with incident events that occurred during an additional 10-year follow-up, adjusting for covariates.

**Results:** We identified four distinct trajectory groups among 6,570 participants (mean age 52; 55% women): Group 1 (n=2,713, reference) consisted of low-level drinkers; Group 2 (n=1,818) mainly included abstainers; Groups 3 (n=805) and 4 (n=1,234) comprised drinkers with varying patterns, with Group 4 consuming the most. Over 10 years of follow-up, women in Groups 2 to 4 had hazard ratios (HR) for CHD of 1.57 (95% CI = 1.11-2.22), 1.57 (95% CI = 1.08-2.29), and 1.27 (95% CI = 1.05-1.55) compared to Group 1. Women in

Groups 2 (HR = 1.26, 95% CI = 1.05-1.50) and 3 (HR = 1.27, 95% CI = 1.05-1.55) also had higher all-cause mortality risks. Men in Group 2 had higher mortality (HR = 1.18, 95% CI = 1.02-1.37, P = 0.030) and CHD (HR = 1.56, 95% CI = 1.07-1.52, P = 0.001) risks compared to Group 1, while men in Group 4 had the highest mortality risk (HR = 1.27, 95% CI = 1.02-1.37).

**Conclusion:** This study identified four distinct alcohol consumption trajectories and demonstrated that maintaining low to moderate levels of alcohol drinking over time is likely associated with a lower risk of CHD and mortality in both women and men.

## INTRODUCTION

Alcohol consumption significantly contributes to the global disease burden and considerably impacts overall health outcomes and mortality rates [1]. Extensive research has explored the effects of alcohol on various health conditions, including organ damage, cognitive decline, and immune deficiency [2-5]. The Global Burden of Diseases study investigated alcohol use in individuals aged 15 to 95 across 195 countries from 1990 to 2016. This study concluded that no minimum amount of alcohol consumption is considered safe [1].

Cardiovascular disease (CVD), including heart disease and stroke, poses a significant health burden in the United States [6]. The relationship between alcohol consumption and CVD is complex and multifaceted [7]. Earlier research suggested that moderate alcohol consumption may have a protective effect against certain types of CVD [8-10]. Recently, methods such as pooled cohort analyses, multivariable-adjusted meta-analyses, and Mendelian randomization have been employed to further explore the relationship between alcohol consumption and CVD [11-14]. These studies suggest that low to moderate alcohol consumption is associated with a higher risk of liver cancer and total mortality and may not offer protective effects against CVD [11-15]. However, most of these studies measured alcohol consumption at a single time point, which may not capture the dynamic nature of an individual’s drinking habits, which may change from time to time due to various life circumstances. For example, research has shown that health-related stressors may lead to changes in alcohol intake over time, with older adults reducing their consumption in response to these stressors [16]. Therefore, investigating long-term alcohol consumption patterns and their relationship with CVD is essential.

This study aims to identify longitudinal alcohol consumption trajectories and to assess whether these trajectory groups have varying risks for coronary heart disease (CHD), the most common form of CVD, and all-cause mortality in the longitudinal Framingham Heart Study (FHS). We hypothesized that distinct patterns of long-term alcohol consumption are associated with different levels of risks for CHD and all-cause mortality. The study results can help target prevention strategies and enhance public health recommendations on alcohol consumption.

## METHOD

### Study participants

Initiated in 1948, the FHS is a community-based, multi-generational, longitudinal cohort study [17, 18]. The present analysis included participants from the Original and Offspring cohorts [17, 18]. Participants in the Original cohort underwent up to 32 exams every 2 to 4 years, and those in the Offspring cohort have completed up to 10 exams every 4 to 8 years. During each exam, comprehensive health and lifestyle information, including alcohol consumption, was collected. We conducted a two-phase study (**Figure 1**): in Phase 1, trajectory analysis was undertaken to classify participants based on their longitudinal alcohol consumption patterns; in Phase 2, we examined the associations of the identified alcohol consumption trajectory groups with CVD risk and mortality.

**Figure 1.**
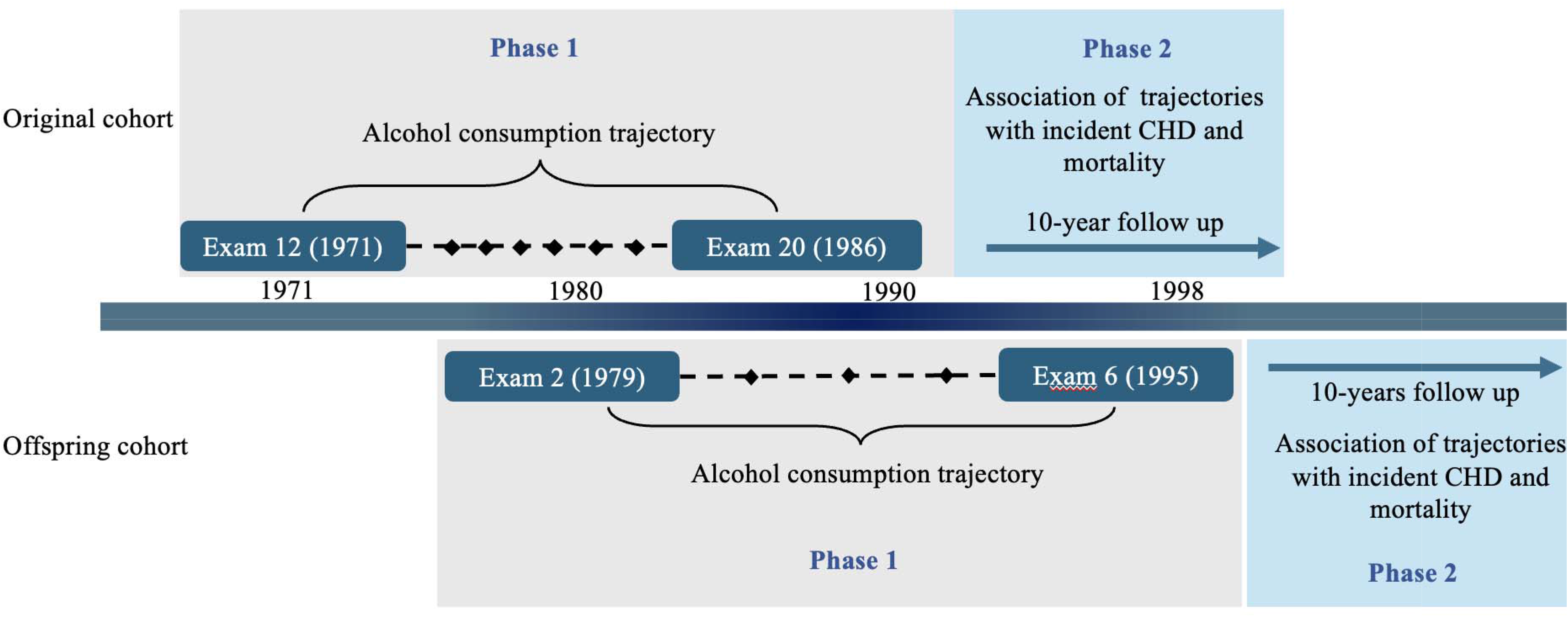
Study timeline and data collection milestones.

For the Original cohort, phase 1 used data from eight exams over 15 years: exam 12 (1971–74, baseline), exams 13, 14, 15, 17, 18, 19, and exam 20 (1986–90). Data collected before Exam 12 was excluded because alcohol consumption data was not regularly collected. Exam 16 data were also excluded due to a high missing rate. For the Offspring cohort, data from five exams were included over 16 years: exam 2 (1979–83, baseline), exams 3, 4, 5, and 6 (1995–98). Exam 1 was excluded due to limited alcohol consumption information.

We also applied several additional exclusion criteria. We excluded those under 18 years of age at baseline (n=6), those with BMI below 18 or above 50 kg/m^2^ (n=82), and those who attended fewer than three exams or with missing alcohol consumption and covariates in two or more consecutive exams (n =713 for the Original cohort and n = 836 for the Offspring cohort). After applying these criteria, 6570 participants were included in the Phase 1 analysis (n = 2790 for the Original cohort, and n = 3780 for the Offspring cohort). In Phase 2, participants with prevalent CVD outcomes at the last attendance in Phase 1 were excluded from further analysis (n = 1545 prevalent CVD and n = 1076 prevalent CHD) in the analysis for incident CHD. For individuals with nonconsecutive missing values, we conducted a K-nearest neighbor imputation model (with k = 2) to impute missing alcohol consumption data and covariates, assuming the missing values followed an intermittent pattern [19, 20] (**Figure 2**).

**Figure 2.**
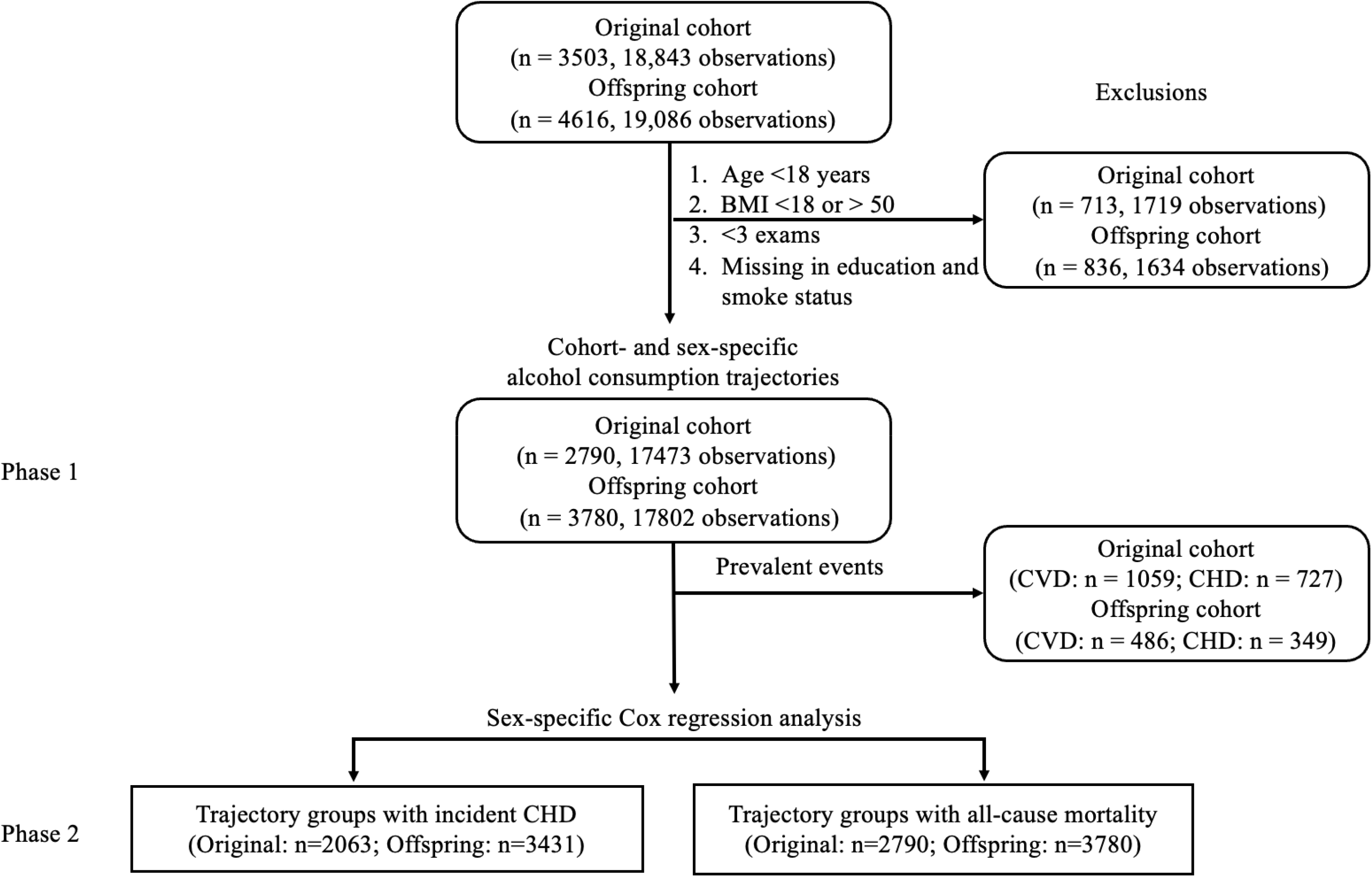
Study design. In Phase 1, trajectory analysis was performed to identify alcohol consumption trajectory groups. In Phase 2, association analysis was performed evaluate if alcohol trajectory groups were associated with CHD and total mortality.

### Alcohol consumption measurement

Alcohol consumption data was collected at each exam through technician-administered questionnaires [21]. Participants reported their average weekly consumption of beer, wine, or liquor (in standard portion size) over the past year. A standard drink was 12 ounces of beer, 4 to 5 ounces of wine, or 1.5 ounces of liquor, each containing approximately 14□grams of ethanol [22]. Given the right-skewed distribution of alcohol consumption, a cohort- and sex-specific Box-Cox transformation was applied to normalize alcohol consumption data [23]. Participants were also classified into four drinking categories based on non-transformed alcohol consumption levels. Non-drinkers were those with zero alcohol consumption (g/day = 0). Moderate drinkers included men who consumed more than 0 but less than 28 g/day of alcohol and women who consumed more than 0 but less than 14 g/day. At-risk drinkers were men with alcohol intake between 28 and 42 g/day and women consuming between 14 and 28 g/day. Participants exceeding these thresholds, with men consuming more than 42 g/day and women consuming more than 28 g/day, were classified as heavy drinkers [22].

### CVD risk score calculation

This study analyzed two risk scores, the FRS (Framingham risk score) and the ASRS (ACC/AHA Atherosclerotic Cardiovascular Disease risk score). These two scores have been widely used for estimating the 10-year risk of developing CVD [24, 25]. The FRS considers factors such as age, sex, smoking status, total cholesterol, high-density lipoprotein cholesterol, systolic blood pressure, and treatment for hypertension [24]. The ASRS includes all FRS variables and incorporates diabetes as a critical factor[25]. The FRS is designed for participants aged 30-79, while the ASRS applies to individuals aged 40-79 [24, 25]. This study calculated both risk scores using data from the last attendance of the Phase 1 study for each participant.

### CHD and all-cause mortality

At FHS, a panel of three physicians adjudicated CHD outcomes and death events by comprehensively reviewing medical records and death certificates [26]. The diagnosis of CHD encompasses coronary insufficiency, recognized myocardial infarction, and CHD death [26, 27].

### Covariates

Covariates included education, current smoking status, systolic blood pressure (SBP), hypertension treatment, diabetes, and BMI at the baseline of Phase 2 [29]. Education was categorized into four levels: no high school, high school, some college, and college graduate. Current smoking status was recorded as a binary variable (Yes or No), indicating whether the participant was smoking at the time of the exam. SBP was measured by physicians, with two readings taken, and the average was used for our analysis [26]. Diabetes was defined as a fasting glucose level equal to or greater than 126 mg/dL or using glucose-lowering medications [30]. Hypertension treatment was recorded based on the use of antihypertensive medications to treat high blood pressure. BMI was calculated by dividing the weight in kilograms by the square of their height in meters (kg/m^2^) for a participant.

### Statistical Analysis

#### Trajectory analysis of alcohol consumption

Growth Mixture Model (GMM) [31] was used to identify longitudinal patterns for alcohol consumption. GMM is a statistical framework combining a growth curve analysis with finite mixture modeling [32], enabling the representation of individual alcohol consumption trajectories as a mixture of latent classes. Each class is characterized by its own growth parameters: initial status, linear growth, and quadratic growth [31, 33]. Covariates in GMM included age, education, and smoking status at baseline in Phase 1. Trajectory models were compared with varying configurations, ranging from one to five trajectory groups [34].

To balance clinical interpretability and model complexity, a second-order polynomial function of time was applied to all five cohort- and sex-specific sample models. The Bayesian Information Criterion (BIC) was used to determine the optimal number of trajectory groups, with the model having the lowest BIC deemed the best fit. We performed 100 iterations for each model and selected the model that yielded the minimum BIC [35]. Additionally, we required each identified trajectory group to contain at least 5% of all study participants to ensure meaningful group size [36, 37].

#### Association analysis of alcohol consumption trajectories with CVD and mortality

Unadjusted one-way ANOVA was used to compare the distribution of FRS and ASRS across trajectory groups to determine if significant differences existed. In Phase 2, cox regression models were used to examine the association between alcohol consumption trajectories and incident CHD and all-cause mortality. For the participants who experienced incident CHD, the follow-up duration was calculated from the baseline of Phase 2 to the date of the first occurrence of CHD. For those without CHD, the follow-up duration was determined from the baseline of Phase 2 to the earliest of the following: last contact date, death, or 10 years after the last attendance of Phase 1. For all-cause mortality, the follow-up time was calculated from baseline to the date of death or the last contact date, whichever occurred first. Covariates included age, sex, BMI, SBP, hypertension treatment, and diabetes. All statistical analyses were performed with R version 4.1.1. A two-sided *P* value of less than 0.05 was considered statistically significant.

## RESULT

### Participant characteristics

At the baseline of Phase 1 (**Figure 1**), we analyzed longitudinal alcohol consumption data from 2790 Original cohort participants (mean age: 63±8, 59.7% women) and 3780 Offspring cohort participants (mean age: 44±10, 52.1% women) (**Table 1**). The Original and Offspring cohorts varied significantly in baseline age and health conditions (*P* < 0.001) (**Table 1**). Half of the Original cohort participants had hypertension (55.7% in women and 50% in men). In contrast, 17% of the women and 28% of men had hypertension in the Offspring cohort. Women in the Original cohort had a larger BMI than those in the Offspring cohort (26.5 vs. 24.7kg/m^2^). In both cohorts, women had comparable alcohol consumption, with a median of 4g/day compared to 2g/day. Similarly, men in both groups had a median alcohol intake of 14g per day (**Table 1**).

**Table 1.** Baseline characteristics of the study samples.

| Variable<sup>1</sup> | Original cohort |  | Offspring cohort |  |
| --- | --- | --- | --- | --- |
|  | Men (N=1123) | Women (N=1667) | Men (N=1812) | Women (N=1968) |
| Age, years | 63 (7) | 64 (8) | 44 (10) | 44 (10) |
| Education, *n* (%) |  |  |  |  |
| No high school | 433 (38.6%) | 611 (36.7%) | 131 (7.2%) | 110 (5.6%) |
| High school | 344 (30.6%) | 546 (32.8%) | 521 (28.8%) | 679 (34.5%) |
| Some college | 148 (13.2%) | 353 (21.2%) | 441 (24.3%) | 635 (32.3%) |
| College or above | 198 (17.6%) | 157 (9.4%) | 719 (39.7%) | 544 (27.6%) |
| BMI, kg/m<sup>2</sup> | 26.94 (3.42) | 26.46 (4.51) | 26.76 (3.59) | 24.66 (4.70) |
| DBP, mmHg | 84.50 (12.14) | 83.63 (12.07) | 82.03 (10.56) | 76.07 (10.61) |
| SBP, mmHg | 141.56 (21.92) | 144.91 (25.73) | 127.37 (17.25) | 119.49 (18.50) |
| TC, mg/dL | 225.07 (38.21) | 227.07 (43.45) | 209.02 (38.01) | 203.85 (40.88) |
| HDL, mg/dL<sup>2</sup> | NA | NA | 43.20 (11.83) | 55.55 (14.87) |
| Hypertension treatment, *n* (%) | 160 (14.3%) | 371 (22.3%) | 194 (10.7%) | 180 (9.1%) |
| Lipid treatment, *n* (%) | 13 (1.2%) | 38 (2.3%) | 21 (1.2%) | 10 (0.5%) |
| Current diabetes, *n* (%) | 4 (9.8%) | 4 (6.6%) | 60 (3.4%) | 36 (1.9%) |
| Obesity, *n* (%) | 176 (15.7%) | 312 (18.8%) | 297 (16.5%) | 238 (12.1%) |
| Hypertension<sup>3</sup>, *n* (%) | 566 (50.4%) | 929 (55.7%) | 506 (27.9%) | 343 (17.4%) |
| Alcohol consumption, g/day<sup>4</sup> | 14 (4, 23) | 2 (0, 12) | 14 (4, 22) | 4 (0, 14) |
| Drinking groups, *n* (%) |  |  |  |  |
| At-risk drinker | 130 (11.6%) | 211 (12.7%) | 191 (10.5%) | 246 (12.5%) |
| Heavy drinker | 194 (17.3%) | 109 (6.5%) | 276 (15.2%) | 134 (6.8%) |
| Moderate drinker | 616 (54.9%) | 795 (47.7%) | 1012 (55.8%) | 1009 (51.3%) |
| Non-drinker | 183 (16.3%) | 552 (33.1%) | 333 (18.4%) | 579 (29.4%) |

<sup>1</sup> Values were presented as mean (SD) for continuous variables and count (percent) for category variables.
<sup>2</sup> HDL information was not available for the Original cohort at exam 12.
<sup>3</sup> Hypertension was defined as the use of hypertension treatment, SBP ≥130 mmHg, or a diastolic blood pressure (DBP) ≥80 mmHg.
<sup>4</sup> Median (IQR) was reported.
Note: All variables showed significant differences across the four groups (all $P < 0.001$ ).

### Identification of alcohol consumption trajectories

We identified five alcohol consumption trajectory groups in both men and women in the Original cohort and Offspring cohorts (**Supplemental Figure 1**). To facilitate a more straightforward interpretation, we combined participants in two trajectory groups among the five with similar trends, resulting in four distinct trajectory groups for the sex and cohort-specific analyses (**Supplemental Figure 2**).

For women, consistent alcohol consumption trajectories were observed across both cohorts (**Supplemental Figure 2)**. Group 1 consisted of moderate drinkers who displayed slightly decreased drinking levels. These women initially consumed approximately 10 g/day and gradually reduced their consumption to less than 5 g/day by the end of Phase 1 (**Supplemental Table 2**). Group 2 consisted of women who maintained minimal alcohol consumption (approximately 0 g/day) throughout the whole period. In Group 3, women started with an average alcohol consumption of ~5 g/day but increased to ~10 g/day around year 7, followed by a gradual decline to 0 g/day by the end of Phase 1. Group 4 included women who consistently consumed alcohol at at-risk levels throughout the 15-year study.

Similarly, consistent drinking patterns were observed for alcohol consumption in men between the two cohorts (**Supplemental Figure 2**). Group 1 consisted of moderate drinkers with a slightly decreasing trend over time. Starting with an average of 20 g/day at the beginning of Phase 1, their consumption reduced to 12 g/day by the end. In Group 2, participants were abstainers (Original cohort) or were abstainers after 10 years of low-level consumption (Offspring cohort). Group 3 consisted of men who started as at-risk drinkers who consumed 30 g/day of alcohol. They increased their consumption to approximately 35– 40 g/day by year 10. However, by the end of Phase 1, these men became non-drinkers in both cohorts. In Group 4, the participants were at risk or heavy drinkers, starting at around 60 g/day at baseline and reducing to 45 g/day by the end of Phase 1 in both cohorts.

The Original and Offspring cohorts exhibited similar alcohol trajectories for their respective groups; thus, we combined the sex-specific trajectory group with similar trajectories from the two cohorts (**Figure 3**). These combined sex-specific trajectory groups were used in the association analyses in Phase 2. Group 1 included moderate drinkers who slightly decreased their consumption. Group 2 included participants who were long-term non-drinkers. Group 3 included participants who experienced a sharp increase in drinking followed by a decrease. Participants in Group 4 were long-term at-risk or heavy drinkers who slightly decreased their drinking.

**Figure 3.**
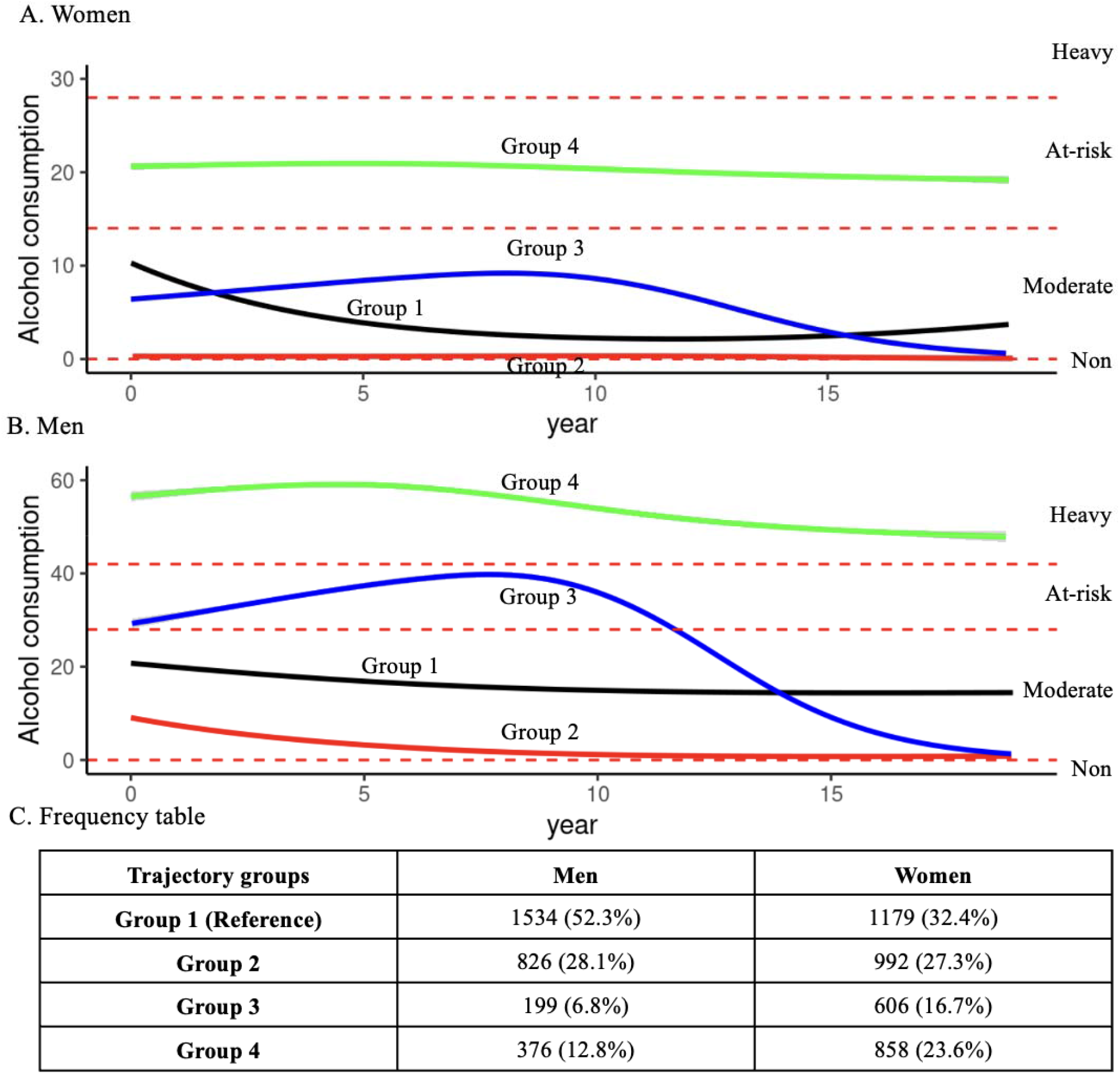
Alcohol consumption trajectory groups in women (A) and men (B). The number and percentage of participants for each trajectory group are included in C. T alcohol trajectory groups fall into four categories based on grams per day of alcohol consumption, including non-drinkers, moderate drinkers (0-14 g/day for women and 28 g/day for men), at-risk drinkers (14-28 g/day for women and 28-42 g/day for men), and heavy drinkers (> 28 g/day for women or > 42 g/day for men). Group 1 includ moderate drinkers who slightly decreased their consumption. Group 2 included participants who were long-term non-drinkers. Group 3 included participants wh experienced a sharp increase in drinking followed by a decrease. Participants in Group 4 were long-term at-risk or heavy drinkers who slightly decreased their drinking.

### Comparison of CVD risk scores among alcohol consumption trajectories

In women, Group 2 has the highest rates of diabetes (12.4%), hypertension (41.8%), and obesity (28.6%) at the Phase 1 baseline (**Supplemental Table 3**). Similarly, men in Group 2 have the highest rates of diabetes and obesity. However, men in Group 4 have the highest rate of hypertension (**Supplemental Table 4**).

Among women, the average CVD risk scores significantly varied among the four alcohol consumption trajectory groups (*P* < 0.001). Those in Group 2 (FRS = 15.5, ASRS = 13.5) and Group 3 (FRS = 15.9, ASRS = 14.4) had higher CVD risk scores, whereas women in Group 4 had the lowest CVD risk scores (FRS = 12.3, ASRS = 10.0) compared to Group 1 (FRS = 14.27, ASRS = 12.58). (**Supplemental Table 2**). The average FRS for men significantly varied among the four alcohol consumption trajectory groups (P = 0.018), while the ASRS did not (P = 0.403). Men in Group 4 had the highest CVD risk scores (FRS = 30.1, ASRS = 20.2). The other three groups of men showed comparable CVD risk scores across both cohorts (**Supplemental Table 3**).

### Association of alcohol consumption trajectories with incident CHD

Among women, 13.6% (n = 493) had incident CVD, and 7.2% (n = 263) developed CHD during a median of 10 years of follow-up. For men, 15.2% (n = 445) developed CVD, and 10.6% (n = 312) developed CHD over the median 10-year follow-up (**Supplemental Table 2**).

Women in both Groups 2 and 3 showed a 57% higher risk of developing CHD compared to those in Group 1 (Group 2: 95% CI = 1.11-2.22, *P* = 0.01; Group 3: 95% CI = 1.08 - 2.29, *P* = 0.02), adjusting for age, BMI, smoking status, hypertension, diabetes, and SBP at the baseline of Phase 2. In Group 4, women had developing CHD compared to Group 1 a 62% higher risk of developing CHD compared to Group after adjusting for covariates (95% CI = 1.11-2.36, *P* = 0.01) (**Figure 4**; **Supplemental Table 9**).

**Figure 4.**
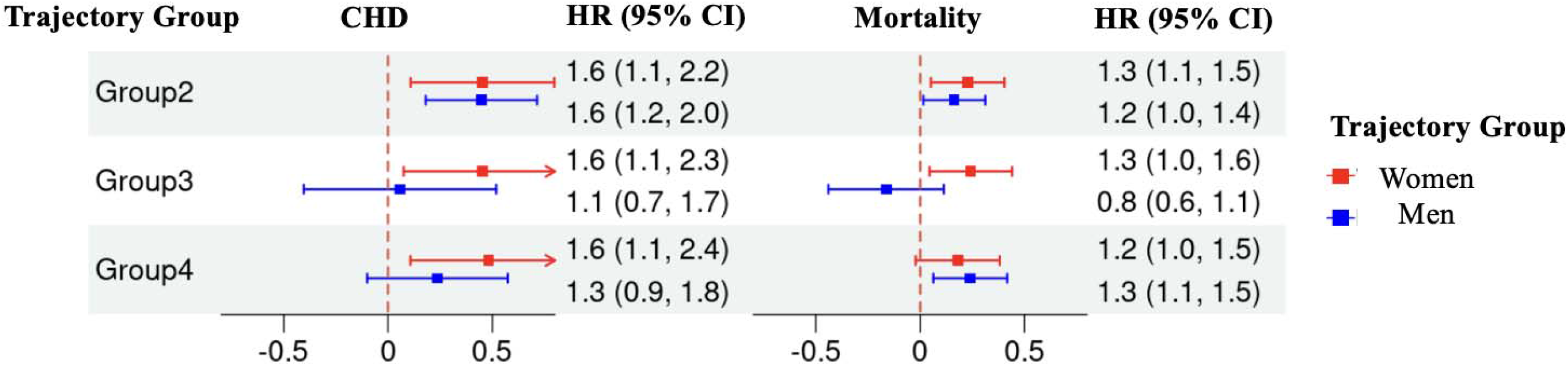
Sex-stratified association analysis between alcohol consumption trajectory groups with CHD and all-cause mortality. The alcohol consumption groups w described in Figure 3. Covariates include age, body mass index, current smoking status, systolic blood pressure, hypertension treatment, and diabetes. HR, hazard ratio. CI, 95% confidence interval.

Men in Group 2 had a 56% higher risk of developing CHD compared to Group 1 after adjusting for covariates (95% CI = 1.20 - 2.04, P = 0.001). However, after adjusting for covariates, men in Groups 3 and 4 did not show a significantly different risk of developing CHD from Group 1. (**Supplemental Table 10**).

### Association of alcohol consumption trajectories with all-cause mortality

Approximately 30.9% of women (1124) and 38.9% of men (1141) died during a median 10-year follow-up period. For women, we found a significantly higher mortality risk in both Group 2 (HR = 1.26, 95% CI = 1.05-1.50, *P* = 0.01) and Group 3 (HR = 1.27, 95% CI = 1.05-1.55, *P* = 0.016) compared to Group 1 after adjusting for covariates, including age, BMI, smoking status, hypertension, diabetes, and SBP at the baseline of Phase 2 (**Figure 4**; **Supplemental Table 5**). Men in Group 2 had a 21% higher hazard of all-cause mortality compared to Group 1 (slightly decreased moderate drinkers) in the base model (95% CI = 1.05-1.39, *P* = 0.008) and 18% higher hazards (95% CI = 1.02-1.37, *P* = 0.03) after adjusting for covariates at the baseline. Similarly, men in Group 4 had a 28% higher hazard of all-cause mortality compared to Group 1 in the base model (95% CI = 1.08-1.52, *P* = 0.005) and a 27% higher risk (95% CI = 1.07-1.52, *P* = 0.008) after adjusting for covariates. No significant associations were observed for men in Group 3 compared to Group 1 (**Figure 4**; **Supplemental Table 6**).

## DISCUSSION

This study identified four distinct, sex-specific alcohol consumption trajectories in participants from two longitudinal cohorts in the FHS. It demonstrated differing associations between these trajectories and incident CHD and all-cause mortality in middle and older adulthood. Most participants displayed an overall decreasing trend in their alcohol consumption across 15 years. However, the pattern of this decline varied across trajectory groups in both women and men. Using Group 1 as a reference, where participants consumed moderate but gradually decreasing amounts of alcohol during the 15-year observation period, we found that women in all other three groups displayed significantly higher hazards for incident CHD and total mortality. For men, compared to those in Group 1, long-term abstainers (Group 2) displayed higher hazards for incident CHD and all-cause mortality, while long-term heavy drinkers (Group 4) had higher hazards for all-cause mortality.

Our study showed different association patterns between trajectory groups and the risk of developing CHD and all-cause mortality in men and women. For example, in the increase and decrease trajectory (Group 3), no significant increase in CHD and all-cause mortality risk was observed in men compared to Group 1. In contrast, women in Group 3 had a higher risk of both mortality and CHD compared to Group 1. These findings underscore the importance of considering sex-specific drinking trajectories in evaluating cardiovascular and overall health.

Our findings support the previous studies showing that stable, moderate alcohol consumption is associated with a lower risk of developing all-cause mortality compared to abstinence [9]. This favorable relationship for moderate drinking has also been reported for some diseases in the 2016 Alcohol Collaborators of Global Burden of Disease, e.g., light alcohol intake is associated with a lower risk of ischemic heart disease and diabetes in women [1]. However, it is worth noting that abstainers tend to have existing health conditions such as obesity, diabetes, and hypertension, which are major risk CVD factors. It is also worth mentioning that alcohol may have adverse impacts on other diseases such as cancer and infectious diseases [1]. Therefore, we should be cautious about adopting moderate drinking habits solely to protect against CHD or promote longevity.

This study has several limitations. Although we considered multiple covariates affecting alcohol consumption patterns in our trajectory modeling, there might be some unmeasured covariates or measurement errors that could bias our findings. Additionally, future research might look at how covariates change over time. We also acknowledge that the self-reported alcohol consumption data may not accurately reflect real drinking habits. Our trajectory analysis encompassed up to seven repeated alcohol measurements, but additional measurements may be necessary to capture the trajectories more accurately. FHS Original and Offspring cohorts are predominantly White individuals of European descent with higher levels of education and socioeconomic status, which may limit the generalizability of our findings to more diverse populations [38]. Therefore, future studies should include a more diverse group of individuals to enhance the applicability of the results across different populations. Despite the limitations, this study has several advantages. First, the 15 years of alcohol consumption data provide a valuable resource for comprehensively quantifying drinking patterns. Additionally, this study included two cohorts with different age demographics, allowing for trajectory analyses across generations.

In summary, this study identified four distinct alcohol consumption trajectories for both men and women across two FHS cohorts. Our findings suggest that maintaining low to moderate levels of alcohol drinking over time is likely associated with a lower risk of CHD and mortality in both women and men. These results emphasize the importance of understanding long-term alcohol consumption patterns in public health interventions.

## Data Availability

The data could be requested through an application to the FHS (https://www.framinghamheartstudy.org/fhs-for-researchers).

## Acknowledgments

We extend our sincere gratitude to the participants of the Framingham Heart Study for their dedication. This research would not have been possible without their invaluable contributions.

