## Supplemental for "Fifteen-year Alcohol Consumption Trajectories and Their Association with Cardiovascular Events and Mortality: the Framingham Heart Study"

**Supplemental Figure 1.** Alcohol consumption trajectory groups in women (A) and men (B). The number and percentage of participants for each trajectory group are included in C. The alcohol trajectory groups fall into four categories based on grams per day of alcohol consumption, including non-drinkers, moderate drinkers (0-14 g/day for women and 0-28 g/day for men), at-risk drinkers (14-28 g/day for women and 28-42 g/day for men), and heavy drinkers (> 28 g/day for women or > 42 g/day for men).

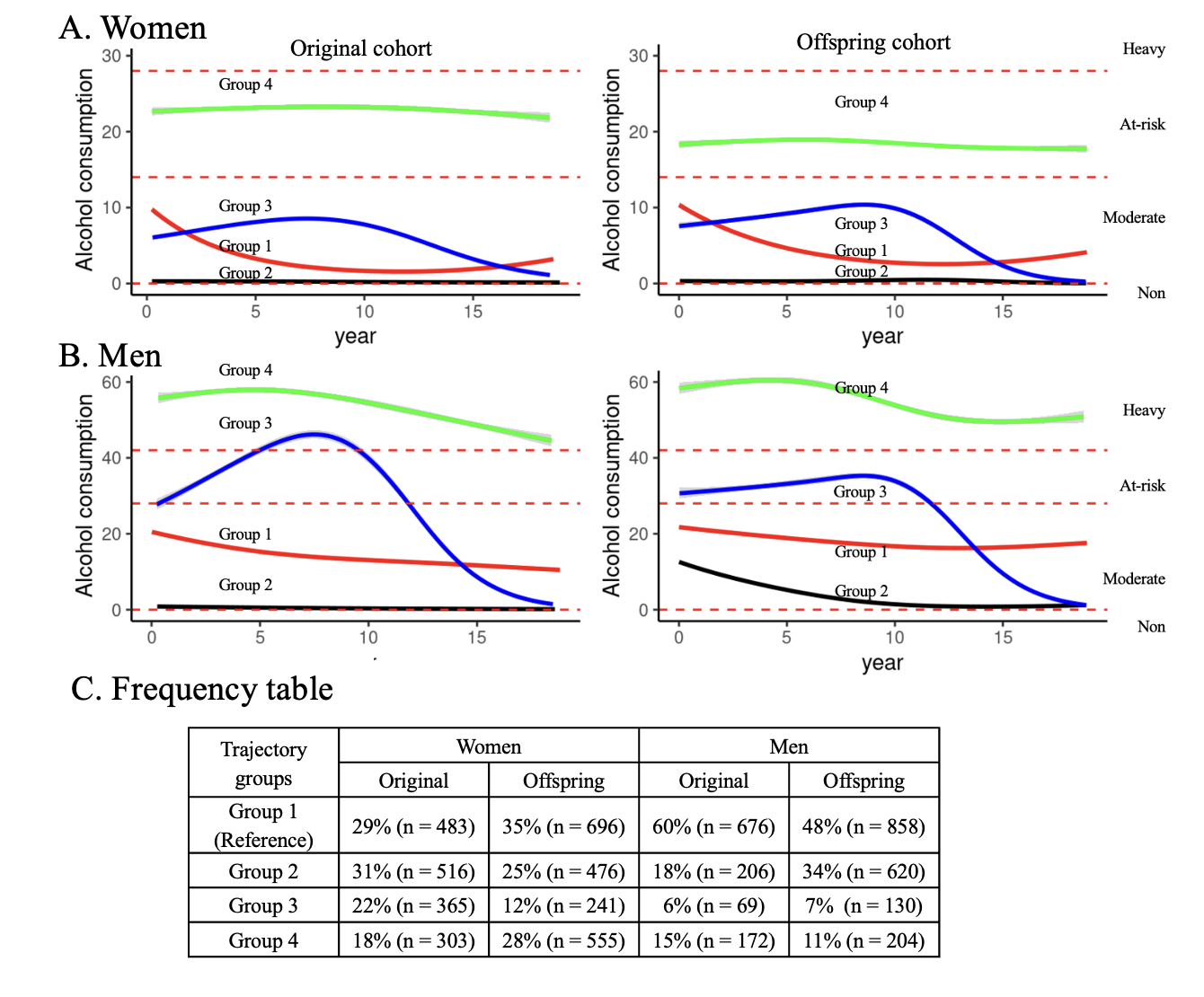

**Supplemental Figure 2.** Alcohol consumption trajectory groups in women (A) and men (B). The number and percentage of participants for each trajectory group are included in C. The alcohol trajectory groups fall into four categories based on grams per day of alcohol consumption, including non-drinkers, moderate drinkers (0-14 g/day for women and 0-28 g/day for men), at-risk drinkers (14-28 g/day for women and 28-42 g/day for men), and heavy drinkers (> 28 g/day for women or > 42 g/day for men).

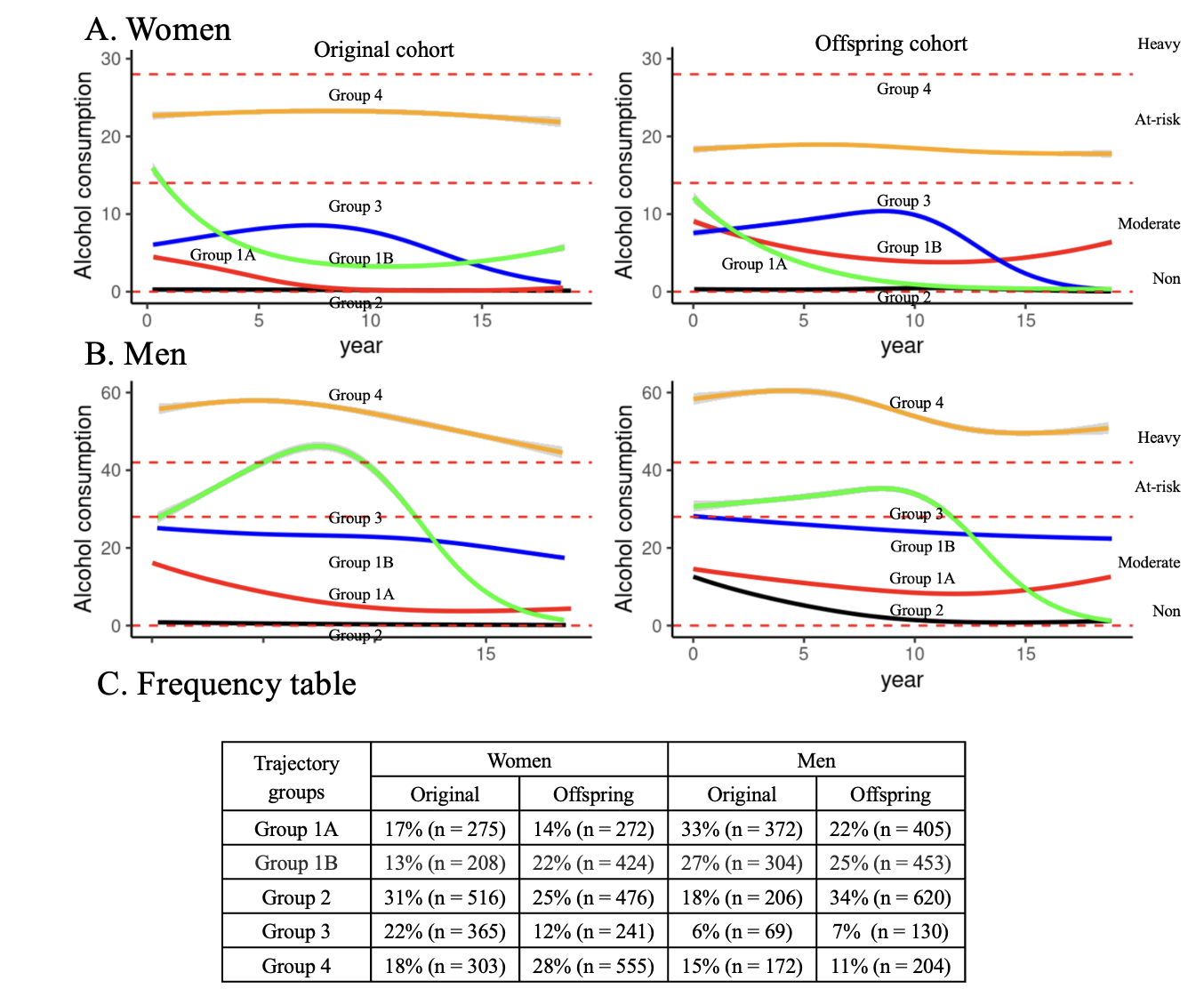

**Supplemental Figure 3.** Unadjusted sex-stratified association analysis between alcohol consumption trajectory groups with CHD and all-cause mortality. The alcohol consumption groups are described in Figure 3. HR, hazard ratio. 95% CI, 95% confidence interval.

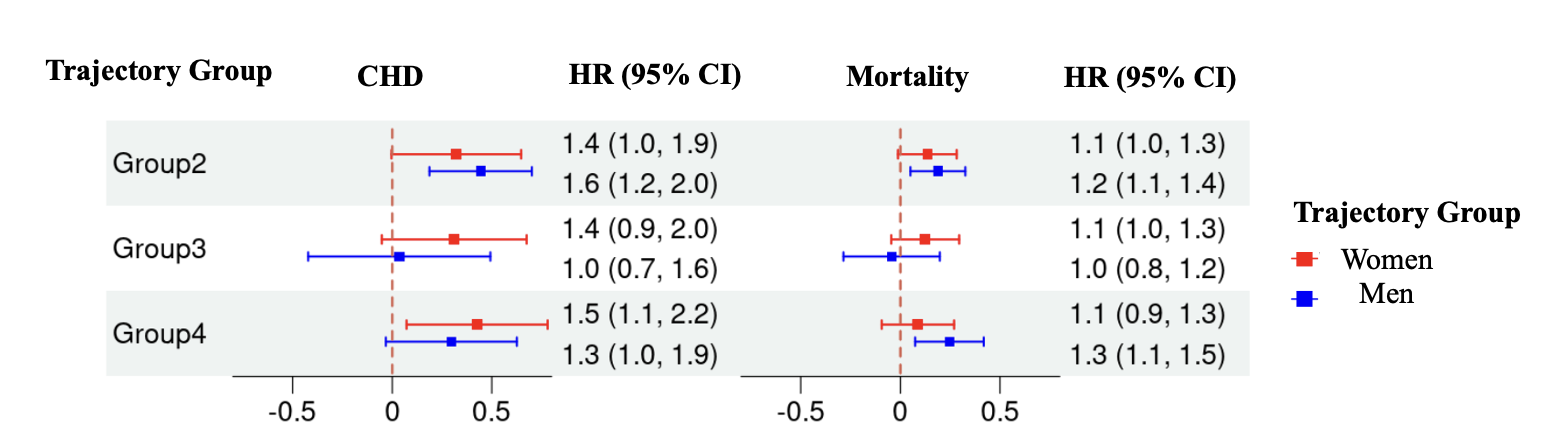

**Supplemental Table 1**. Characteristics of the study samples in Phase 2.

| **Variable**^1^ | **Men (n=2935)** | **Women (n=3635)** | ***P* value** |
| --- | --- | --- | --- |
| Age, years | 65 (12) | 67 (13) | < 0.001 |
| Education, *n* (%) |  |  | < 0.001 |
| No high school | 564 (19.2%) | 721 (19.8%) |  |
| High school | 865 (29.5%) | 1225 (33.7%) |  |
| Some college | 589 (20.1%) | 988 (27.2%) |  |
| College or above | 917 (31.2%) | 701 (19.3%) |  |
| BMI, kg/m^2^ | 27.68 (4.22) | 26.74 (5.17) | < 0.001 |
| DBP, mmHg | 80.56 (11.67) | 77.67 (11.83) | < 0.001 |
| SBP, mmHg | 140.28 (22.83) | 140.96 (26.67) | 0.277 |
| TC, mg/dL | 201.28 (40.41) | 216.08 (40.65) | < 0.001 |
| HDL, mg/dL^2^ | 42.80 (12.52) | 55.56 (16.14) | < 0.001 |
| Hypertension treatment, *n* (%) | 1021 (35.0%) | 1311 (36.2%) | 0.345 |
| Lipid treatment, *n* (%) | 298 (10.2%) | 241 (6.6%) | < 0.001 |
| Current diabetes, *n* (%) | 361 (12.6%) | 291 (8.4%) | < 0.001 |
| Obesity, *n* (%) | 699 (24.6%) | 738 (21.9%) | 0.011 |
| Hypertension^3^, *n* (%) | 1588 (54.1%) | 1969 (54.2%) | 0.95 |
| Alcohol consumption, g/day^4^ | 6 (0, 22) | 0 (0, 8) | < 0.001 |
| Drinking groups^1^ *n* (%) |  |  | < 0.001 |
| At-risk drinker | 251 (8.6%) | 312 (8.6%) |  |
| Heavy drinker | 253 (8.6%) | 156 (4.3%) |  |
| Moderate drinker | 1416 (48.2%) | 1284 (35.3%) |  |
| Non-drinker | 1015 (34.6%) | 1883 (51.8%) |  |
| FRS | 27.35 (19.43) | 13.97 (13.85) | < 0.001 |
| ASRS | 19.34 (15.23) | 11.99 (12.59) | < 0.001 |
| Incident CVD, *n* (%) | 445 (15.2%) | 493 (13.6%) | 0.065 |
| Incident CHD, *n* (%) | 312 (10.6%) | 263 (7.2%) | < 0.001 |
| All-cause mortality, *n* (%) | 1141 (38.9%) | 1124 (30.9%) | < 0.001 |
| Mortality follow up, years^4^ | 10 (4.27, 10) | 10 (7.32, 10) | < 0.001 |
| CVD follow up, years^4^ | 10 (4.83, 10) | 10 (6.86, 10) | < 0.001 |
| CHD follow up, years^4^ | 10 (4.78, 10) | 10 (7.24, 10) | < 0.001 |

^1^Values were presented as mean (SD) for continuous variables and count (percent) for category variables.

^2^HDL information was not available for Original cohort at exam 12.

^3^Hypertension was defined as the use of hypertension treatment, SBP ≥130 mmHg, or diastolic blood pressure (DBP) ≥80 mmHg.

^4^Median (IQR) was reported.

**Supplemental Table 2:** median alcohol consumption for trajectory groups (baseline, end)

| Group | Original Cohort | | Offspring Cohort | |
| --- | --- | --- | --- | --- |
|  | Women | Men | Women | Men |
| 1 | (2, 2) | (28, 0) | (4, 8) | (24, 18) |
| 2 | (0, 0) | (0, 4) | (0, 0) | (4, 0) |
| 3 | (0, 0) | (12, 0) | (0, 0) | (2, 0) |
| 4 | (34, 16) | (16, 58) | (16, 32) | (64, 72) |

**Supplemental Table 3.** Characteristics of the study sample in Phase 2 stratified by trajectory groups for women

| **Variable^1^** | **Trajectory group** | | | | |
| --- | --- | --- | --- | --- | --- |
|  | **1 (n=1179)** | **2 (n=992)** | **3 (n=606)** | **4 (n=858)** | ***P* value** |
| Age, years | 66 (13) | 69 (13) | 69 (12) | 63 (12) | < 0.001 |
| Education, *n* (%) |  |  |  |  | < 0.001 |
| No high school | 216 (18.3%) | 280 (28.2%) | 137 (22.6%) | 88 (10.3%) |  |
| High school | 404 (34.3%) | 356 (35.9%) | 204 (33.7%) | 261 (30.4%) |  |
| Some college | 330 (28.0%) | 244 (24.6%) | 166 (27.4%) | 248 (28.9%) |  |
| College or above | 229 (19.4%) | 112 (11.3%) | 99 (16.3%) | 261 (30.4%) |  |
| BMI, kg/m^2^ | 26.99 (5.08) | 27.66 (5.60) | 26.69 (5.20) | 25.52 (4.55) | < 0.001 |
| DBP, mmHg | 77.25 (11.70) | 77.84 (12.18) | 78.35 (11.95) | 77.55 (11.49) | 0.284 |
| SBP, mmHg | 139.49 (26.99) | 143.86 (26.34) | 143.80 (27.37) | 137.60 (25.57) | < 0.001 |
| TC, mg/dL | 216.42 (40.51) | 214.20 (40.45) | 216.46 (41.64) | 217.30 (40.43) | 0.493 |
| HDL, mg/dL^2^ | 54.86 (14.99) | 50.26 (13.73) | 54.00 (15.28) | 62.54 (17.78) | < 0.001 |
| Hypertension treatment, *n* (%) | 401 (34.2%) | 413 (41.8%) | 245 (40.5%) | 252 (29.4%) | < 0.001 |
| Lipid treatment, *n* (%) | 89 (7.6%) | 71 (7.2%) | 36 (5.9%) | 45 (5.2%) | 0.156 |
| Current diabetes, *n* (%) | 101 (8.9%) | 113 (12.4%) | 42 (7.4%) | 35 (4.2%) | < 0.001 |
| Obesity, *n* (%) | 244 (22.1%) | 248 (28.6%) | 121 (21.6%) | 125 (14.9%) | < 0.001 |
| Hypertension^3^, *n* (%) | 606 (51.4%) | 602 (60.7%) | 358 (59.1%) | 403 (47.0%) | < 0.001 |
| Alcohol consumption, g/day^4^ | 2 (0, 4) | 0 (0, 0) | 0 (0, 2) | 16 (10, 38) | < 0.001 |
| Drinking groups^1^ *n* (%) |  |  |  |  | < 0.001 |
| At-risk drinker | 23 (2.0%) | 0 (0.0%) | 11 (1.8%) | 278 (32.4%) |  |
| Heavy drinker | 1 (0.1%) | 0 (0.0%) | 1 (0.2%) | 154 (17.9%) |  |
| Moderate drinker | 614 (52.1%) | 52 (5.2%) | 193 (31.8%) | 425 (49.5%) |  |
| Non-drinker | 541 (45.9%) | 940 (94.8%) | 401 (66.2%) | 1 (0.1%) |  |
| FRS | 13.46 (14.27) | 15.47 (14.24) | 15.90 (14.65) | 12.34 (12.26) | < 0.001 |
| ASRS | 11.44 (12.58) | 13.45 (13.07) | 14.37 (13.69) | 10.04 (11.07) | < 0.001 |
| Incident CVD, *n* (%) | 132 (11.2%) | 105 (10.6%) | 61 (10.1%) | 112 (13.1%) | 0.253 |
| Incident CHD, *n* (%) | 50 (4.2%) | 44 (4.4%) | 27 (4.5%) | 45 (5.2%) | 0.741 |
| All-cause mortality, *n* (%) | 372 (31.6%) | 307 (30.9%) | 227 (37.5%) | 293 (34.1%) | 0.030 |
| Mortality follow up, years^4^ | 10 (9.03, 10) | 10 (4.45, 10) | 10 (5.59, 10) | 10 (10, 10) | < 0.001 |
| CVD follow up, years^4^ | 10 (8.31, 10) | 10 (4.77, 10) | 10 (4.87, 10) | 10 (9.43, 10) | < 0.001 |
| CHD follow up, years^4^ | 10 (9.55, 10) | 10 (4.69, 10) | 10 (5.39, 10) | 10 (10, 10) | < 0.001 |

^1^Values were presented as mean (SD) for continuous variables and count (percent) for category variables.

^2^HDL information was not available for the Original cohort at exam 12.

^3^Hypertension was defined as the use of hypertension treatment, SBP ≥130 mmHg, or diastolic blood pressure (DBP) ≥80 mmHg.

^4^Median (IQR) was reported.

**Supplemental Table 4**. Characteristics of the study sample in Phase 2 stratified by trajectory groups for men

| **Variable^1^** | **Trajectory group** | | | | |
| --- | --- | --- | --- | --- | --- |
|  | **1 (n=1534)** | **2 (n=826)** | **3 (n=199)** | **4 (n=376)** | ***P* value** |
| Age, years | 65 (12.10) | 64 (12) | 65 (12) | 66 (11) | 0.006 |
| Education, *n* (%) |  |  |  |  | 0.212 |
| No high school | 287 (18.7%) | 160 (19.4%) | 36 (18.1%) | 81 (21.5%) |  |
| High school | 423 (27.6%) | 265 (32.1%) | 61 (30.7%) | 116 (30.9%) |  |
| Some college | 313 (20.4%) | 165 (20.0%) | 45 (22.6%) | 66 (17.6%) |  |
| College or above | 511 (33.3%) | 236 (28.6%) | 57 (28.6%) | 113 (30.1%) |  |
| BMI, kg/m^2^ | 27.54 (4.15) | 28.04 (4.51) | 27.76 (4.12) | 27.42 (3.89) | 0.033 |
| DBP, mmHg | 80.64 (11.94) | 79.58 (11.06) | 79.14 (10.93) | 83.14 (11.84) | < 0.001 |
| SBP, mmHg | 140.68 (23.85) | 137.81 (21.20) | 137.07 (20.62) | 145.76 (22.12) | < 0.001 |
| TC, mg/dL | 202.63 (37.28) | 196.75 (39.63) | 195.42 (34.76) | 209.28 (53.47) | < 0.001 |
| HDL, mg/dL^2^ | 43.64 (12.51) | 39.00 (10.86) | 42.31 (11.52) | 48.68 (13.91) | < 0.001 |
| Hypertension treatment, *n* (%) | 495 (32.5%) | 294 (35.8%) | 74 (37.4%) | 158 (42.5%) | 0.003 |
| Lipid treatment, *n* (%) | 144 (9.4%) | 96 (11.6%) | 19 (9.6%) | 39 (10.4%) | 0.403 |
| Current diabetes, *n* (%) | 149 (10.0%) | 146 (18.0%) | 27 (14.4%) | 39 (10.6%) | < 0.001 |
| Obesity, *n* (%) | 348 (23.5%) | 214 (26.8%) | 47 (25.8%) | 90 (24.3%) | 0.351 |
| Hypertension^3^, *n* (%) | 815 (53.1%) | 435 (52.7%) | 99 (49.7%) | 239 (63.6%) | < 0.001 |
| Alcohol consumption, g/day^4^ | 12 (4, 20) | 0 (0, 0) | 0 (0, 2) | 44 (34, 64) | < 0.001 |
| Drinking groups^1^ *n* (%) |  |  |  |  | < 0.001 |
| At-risk drinker | 135 (8.8%) | 0 (0.0%) | 4 (2.0%) | 112 (29.8%) |  |
| Heavy drinker | 50 (3.3%) | 0 (0.0%) | 6 (3.0%) | 197 (52.4%) |  |
| Moderate drinker | 1145 (74.6%) | 158 (19.1%) | 47 (23.6%) | 66 (17.6%) |  |
| Non-drinker | 204 (13.3%) | 668 (80.9%) | 142 (71.4%) | 1 (0.3%) |  |
| FRS | 26.16 (18.92) | 28.23 (20.02) | 27.14 (18.56) | 30.07 (20.18) | 0.018 |
| ASRS | 19.02 (15.00) | 19.16 (15.66) | 20.07 (15.39) | 20.68 (15.02) | 0.403 |
| Incident CVD, *n* (%) | 132 (8.6%) | 72 (8.7%) | 22 (11.1%) | 38 (10.1%) | 0.580 |
| Incident CHD, *n* (%) | 57 (3.7%) | 36 (4.4%) | 11 (5.5%) | 17 (4.5%) | 0.593 |
| All-cause mortality, *n* (%) | 436 (28.4%) | 201 (24.3%) | 62 (31.2%) | 114 (30.3%) | 0.056 |
| Mortality follow up, years^4^ | 10 (4.33, 10) | 10 (4.77, 10) | 10 (6.2, 10) | 10 (3.27, 10) | 0.080 |
| CVD follow up, years^4^ | 10 (5.2, 10) | 10 (4.86, 10) | 10 (5.89, 10) | 10 (3.12, 10) | 0.024 |
| CHD follow up, years^4^ | 10 (5.27, 10) | 10 (4.81, 10) | 10 (5.2, 10) | 10 (3.36, 10) | 0.022 |

^1^Values were presented as mean (SD) for continuous variables and count (percent) for category variables.

^2^HDL information was not available for the Original cohort at exam 12.

^3^Hypertension was defined as the use of hypertension treatment, SBP ≥130 mmHg, or diastolic blood pressure (DBP) ≥80 mmHg.

^4^Median (IQR) was reported.

### Supplemental Table 5. Association between alcohol consumption trajectories and all-cause mortality in women.

| **Model** | **Group** | **Beta** | **HR** | **95% CI** | ***P* value** |
| --- | --- | --- | --- | --- | --- |
| Multi-adjusted^*^ | 2 | 0.23 | 1.26 | (1.05, 1.50) | **0.0113** |
|  | 3 | 0.24 | 1.27 | (1.05, 1.55) | **0.0160** |
|  | 4 | 0.18 | 1.20 | (0.98, 1.46) | 0.0802 |
| Crude | 2 | 0.14 | 1.15 | (0.99, 1.33) | 0.0715 |
|  | 3 | 0.12 | 1.13 | (0.96, 1.34) | 0.1517 |
|  | 4 | 0.09 | 1.09 | (0.91, 1.31) | 0.3439 |

*Covariates include education, current smoking status, SBP, hypertension treatment, diabetes, and BMI.

### Supplemental Table 6. Association between alcohol consumption trajectories and all-cause mortality for men.

| **Model** | **Group** | **Beta** | **HR** | **95% CI** | ***P* value** |
| --- | --- | --- | --- | --- | --- |
| Multi-adjusted^*^ | 2 | 0.16 | 1.18 | (1.02, 1.37) | **0.0303** |
|  | 3 | -0.16 | 0.85 | (0.64, 1.12) | 0.2467 |
|  | 4 | 0.24 | 1.27 | (1.07, 1.52) | **0.0078** |
| Crude | 2 | 0.19 | 1.21 | (1.05, 1.39) | **0.0075** |
|  | 3 | -0.04 | 0.96 | (0.75, 1.22) | 0.7232 |
|  | 4 | 0.25 | 1.28 | (1.08, 1.52) | **0.0051** |

*Covariates include education, current smoking status, SBP, hypertension treatment, diabetes, and BMI.

### Supplemental Table 7. Association between alcohol consumption trajectories and CVD for women.

| **Model** | **Group** | **Beta** | **HR** | **95% CI** | ***P* value** |
| --- | --- | --- | --- | --- | --- |
| Multi-adjusted^*^ | 2 | 0.10 | 1.10 | (0.86, 1.41) | 0.4276 |
|  | 3 | 0.12 | 1.12 | (0.86, 1.48) | 0.3949 |
|  | 4 | 0.24 | 1.27 | (0.98, 1.65) | 0.0742 |
| Crude | 2 | 0.00 | 1.00 | (0.79, 1.26) | 0.9897 |
|  | 3 | 0.01 | 1.01 | (0.78, 1.31) | 0.9630 |
|  | 4 | 0.17 | 1.18 | (0.92, 1.51) | 0.1849 |

*Covariates include education, current smoking status, SBP, hypertension treatment, diabetes, and BMI.

### Supplemental Table 8. Association between trajectories of alcohol consumption trajectories and CVD for men.

| **Model** | **Group** | **Beta** | **HR** | **95%CI** | ***P* value** |
| --- | --- | --- | --- | --- | --- |
| Multi-adjusted^*^ | 2 | 0.23 | 1.26 | (1.01, 1.59) | **0.0444** |
|  | 3 | -0.12 | 0.89 | (0.6, 1.31) | 0.5519 |
|  | 4 | 0.04 | 1.04 | (0.78, 1.39) | 0.7892 |
| Crude | 2 | 0.21 | 1.23 | (0.98, 1.53) | 0.0689 |
|  | 3 | -0.04 | 0.96 | (0.66, 1.38) | 0.8145 |
|  | 4 | 0.09 | 1.10 | (0.83, 1.46) | 0.5274 |

*Covariates include education, current smoking status, SBP, hypertension treatment, diabetes, and BMI.

#### Supplemental Table 9. Association between alcohol consumption trajectories and CHD for women.

| **Model** | **Group** | **Beta** | **HR** | **95% CI** | ***P* value** |
| --- | --- | --- | --- | --- | --- |
| Multi-adjusted^*^ | 2 | 0.45 | 1.57 | (1.11, 2.22) | **0.0100** |
|  | 3 | 0.45 | 1.57 | (1.08, 2.29) | **0.0188** |
|  | 4 | 0.48 | 1.62 | (1.11, 2.36) | **0.0117** |
| Crude | 2 | 0.32 | 1.38 | (1.00, 1.91) | 0.0529 |
|  | 3 | 0.31 | 1.36 | (0.95, 1.96) | 0.0935 |
|  | 4 | 0.43 | 1.53 | (1.07, 2.18) | **0.0184** |

*Covariates include education, current smoking status, SBP, hypertension treatment, diabetes, and BMI.

#### Supplemental Table 10. Association between alcohol consumption trajectories and CHD for men.

| **Model** | **Group** | **Beta** | **HR** | **95% CI** | ***P* value** |
| --- | --- | --- | --- | --- | --- |
| Multi-adjusted^*^ | 2 | 0.45 | 1.56 | (1.20, 2.04) | **0.0010** |
|  | 3 | 0.06 | 1.06 | (0.67, 1.68) | 0.8077 |
|  | 4 | 0.24 | 1.27 | (0.9, 1.78) | 0.1682 |
| Crude | 2 | 0.44 | 1.56 | (1.21, 2.02) | **0.0007** |
|  | 3 | 0.04 | 1.04 | (0.66, 1.64) | 0.8807 |
|  | 4 | 0.30 | 1.35 | (0.97, 1.87) | 0.0768 |

^*^Covariates include education, current smoking status, SBP, hypertension treatment, diabetes, and BMI.
